# Burden of Cardiovascular Disease in Brazil, 1996–2023: A Retrospective Descriptive Study of the Epidemiology and Impact on Public Healthcare with Emphasis on Acute Myocardial Infarction

**DOI:** 10.64898/2026.06.19.26356087

**Authors:** Mellyely Carolina Basílio-Soares, João Sérgio Fonseca Guimarães, Ana Luíza Matos de Oliveira, André Gustavo Oliveira, Thiago Verano-Braga

## Abstract

**Background:** Cardiovascular diseases (CVD) are the leading cause of death worldwide, and their epidemiology is correlated with genetic predisposition, exposure to risk factors, sex, age, access to medical care, and other sociodemographic characteristics. Brazil is a developing country with a vast territory, which leads to structural inequalities. Estimates of CVD in Brazil, in its regions, and in its population are poorly evaluated and analysed.

**Methods:** We obtained CVD-related data from the Brazilian Unified Health System (SUS) and analysed mortality and morbidity from 1996 to 2023 by sex, race/ethnicity, age, and region. We calculated the risk of death from the most prevalent diseases, the average length of hospital stay, and the costs associated with heart transplantation.

**Findings:** In Brazil, acute myocardial infarction was the pathology that led to the highest number of deaths across all variables analysed during the evaluated period. Other CVD were also related to causes of death and morbidity, such as hypertensive diseases and heart failure.

**Interpretation:** Brazil presents a serious challenge to the public health system due to the high number of deaths and the progressive mortality rate. This study represents a fundamental contribution to the basis for formulating public health policies aimed at reducing the growing impact associated with these diseases.

**Funding:** CNPq, CAPES, FAPEMIG, INCT

**Research in context:** *Evidence before this study:* Data published annually by the World Health Organization shows that cardiovascular diseases (CVD) are the leading cause of death worldwide, with acute myocardial infarction and stroke being the most prominent. Therefore, we began searching Google Scholar for epidemiological data on CVD over the last 5 years (2021-2025). We used search terms such as “myocardial infarction”, “hypertensive diseases”, and “hospital costs for cardiovascular diseases”. We noted that the studies were scattered in their information, in many cases failing to correlate important parameters for the manifestation of CVD or to measure the proportion of deaths over time. Although the population is aware that CVD are the leading cause of mortality, data available from health systems and records are not explicit and easily interpreted.

*Added value of this study:* This study aimed to evaluate different variables such as hospitalization costs, region, sex, age, and race/ethnicity in relation to the burden of CVD as a percentage of total deaths per 100,000 inhabitants in Brazil. We were able to identify the main causes of mortality from CVD, focusing our study on seven prominent diseases. We confirmed that acute myocardial infarction is still the CVD that causes the most deaths in the country. We analysed sociodemographic parameters with the manifestation of these diseases.

*Implications of all available evidence:* CVD continue to be the leading cause of death in Brazil and worldwide. This study serves as a basis for public health policy development, such as preventive measures. It highlights that the manifestation, incidence, and progression of these diseases are derived from a diverse set of factors, involving social, economic, and geographic spheres.

## Introduction

According to data from the World Health Organization (WHO), cardiovascular diseases (CVD) remain the leading cause of global mortality, accounting for approximately 19.8 million deaths in 2022. CVD arise from a complex interplay of genetic, environmental, and socioeconomic determinants, with modifiable risk factors such as obesity, unhealthy dietary habits, psychosocial stress, physical inactivity, and tobacco use playing a major role in their development and progression ^1–3^.

In a global analysis, data from the Global Health Data Exchange (GHDx), used to estimate the Global Burden of Disease (GBD) study, indicate that CVD-related mortality increased by approximately 18.5% between 1990 and 2023. In Latin America and the Caribbean, the corresponding increase was 8.33%, whereas Brazil experienced an even greater rise of 11.53%. These findings underscore the growing burden of CVD over recent decades, likely driven by population aging, demographic and epidemiological transitions, lifestyle changes, and persistent socioeconomic and healthcare disparities.

Within the International Classification of Diseases (ICD), the ICD-10 version places CVD in Chapter IX. This coding system standardizes clinical and epidemiological records, ensuring greater consistency in data collection and analysis. Identifying the population groups most affected is crucial for shaping public health policies and for designing effective strategies aimed at prevention and the promotion of cardiovascular health.

Among CVD, ischemic heart disease (I25), hypertensive diseases (I10-I15), chronic rheumatic heart disease (I05-I09), acute myocardial infarction (I21), cardiomyopathies (I42), and heart failure (I50) stand out, as they represent some of the most prevalent conditions and are among the leading causes of morbidity and mortality worldwide ^4–6^. The high prevalence of these CVD imposes a substantial economic burden on healthcare systems, increasing both public and private expenditures through greater demand for hospitalizations, diagnostic procedures, chronic pharmacological therapy, and, in advanced cases, high-complexity interventions such as heart transplantation^7^.

In Brazil, health information is managed through the Department of Informatics of the Unified Health System (DATASUS), the official information system of the Brazilian Unified Health System (SUS), established in 1991 to integrate and disseminate national health data. Despite its comprehensive database, DATASUS still presents limitations in data visualization and the integration of multiple variables, hindering more comprehensive epidemiological analyses. In this study, we transformed these large-scale datasets into epidemiological and graphical representations spanning 1996–2023, enabling the investigation of regional, ethnic, sex-, and age-related patterns of cardiovascular disease mortality and morbidity, as well as their economic burden on the Brazilian public healthcare system.

## Methods

### Study design and data collection

All data for this study were retrieved from the Hospital Information System of the Unified Health System (SIH/SUS), managed by the Brazilian Ministry of Health, in collaboration with the Department of Health Care, as well as the State and Municipal Health Departments. All diseases included in this study are classified according to the ICD-10, and are listed under the Chapter IX: Diseases of the Circulatory System. These diseases are described in Table S1, while the analysed variables and parameters are defined in Tables S2 and S3. The tables can be visualised in the supplemental material.

The mortality data is consolidated by the Department of Health Situation Analysis of the Health Surveillance Secretariat, together with the State and Municipal Health Departments. Death certificates are collected from local registry offices, and this data is consolidated and made available through the Mortality Information System; thus, mortality data encompass the entire Brazilian population. It is important to note that, in accordance with rules established by the WHO, the certifying physician is responsible for assigning the cause of death for each individual. Mortality data were analysed from 1996 to 2023, corresponding to the period during which these diseases were classified according to the ICD-10 coding system.

Morbidity data is sent from the hospital units participating with the SUS to municipal and state health authorities and then consolidated by the DATASUS, allowing access to a database that contains hospital admissions carried out in Brazil to the population served by SUS (approximately 70% of the total population). Admission data were retrieved using ICD-10 Chapter IX codes (Table S4, supplemental material) to avoid noise from other diseases. We analysed admissions, relative rates, average stay, and costs from 1998–2023. However, data are not individualized, preventing patient follow-up, distinction between readmissions and new cases, or tracking comorbidities.

The regional divisions used for this study are legally valid as proposed by the Brazilian Institute of Geography and Statistics (IBGE), based on geographical, demographic and economic parameters. This division has five regions of Brazil: North, Northeast, Midwest, Southeast and South. Rates per 100,000 people were calculated with population numbers from censuses or estimates by IBGE. Age standardisation was carried out using the WHO World Standard Population from 2000-2025 ^8^.

### Analysis

Data analysis was carried out through R (version 4.4.2) and RStudio (2024.04.2 Build 764). To model mortality rates associated with CVD, we employed a Jointpoint regression model using the package “segmented” ^9^. This analysis fits a general linear model (GLM) to allow the consideration of piecewise linear relationships. We used Davies’ test for differences in slope to investigate a possible difference in the dynamics of mortality rate over the analysed period.

To calculate age-specific rates of death and age-standardized number of deaths by CVD, we stratified the data into age groups according to the Brazilian Ministry of Health classification: 0–4, 5–9, 10–14, 15–19, 20–29, 30–39, 40–49, 50–59, 60–69, 70–79 and > 80 years old ^10^. Obtained age-specific rates were then weighted by their respective weights according to the WHO world standard population (2000–2025) sizes to get the resulting age-standardised rates.

## Results

### Mortality and morbidity from cardiovascular diseases in Brazil by region, type of heart disease, sex, race and ethnicity, and age

We evaluated the evolution of cardiovascular diseases (CVD) in Brazil between 1996 and 2023 using DATASUS data. Registered deaths peaked in 2022 at approximately 195 per 100,000 inhabitants (Figure 1A), likely reflecting the impact of the COVID-19 pandemic, which was associated with underreporting, worsening CVD outcomes, increased sedentary behaviour, poorer dietary habits, reduced healthcare utilization, and interruptions to ongoing treatments ^11^. In contrast, age-standardized mortality rates declined between 2000 and 2023 (Figure 1A, inset).

**Figure 1.**
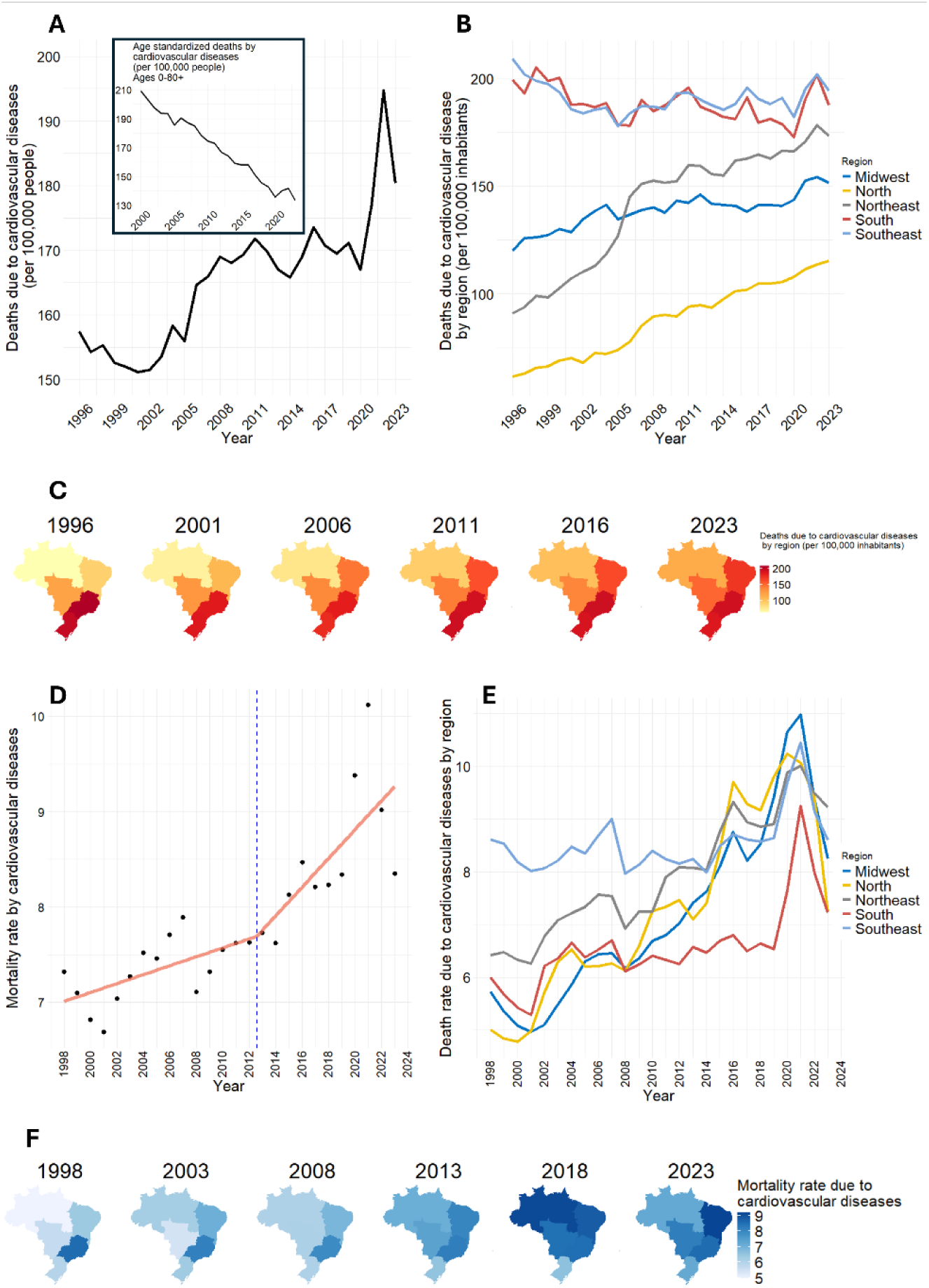
Cardiovascular disease (CVD) mortality in Brazil from 1996 to 2023. (**a**) Absolute number of CVD deaths and crude mortality rate (per 100,000 inhabitants) obtained annually from DATASUS. Inset: age-standardised mortality rate from 2000 to 2023. (**b**) Annual number of CVD deaths according to the Brazilian five geographic regions Midwest, Northeast, North, Southeast, and South. (**c**) Geographic distribution of CVD deaths across Brazil, with colour intensity representing the number of deaths per 100,000 inhabitants. (**d**) Joinpoint regression analysis of the CVD mortality rate from 1998 to 2023, showing the observed values and fitted temporal trends. (**e**) Temporal evolution of CVD mortality rates according to Brazil’s geographic regions. (**f**) Geographic distribution of CVD mortality rates across Brazil at selected time points, with colour intensity representing the mortality rate.

From the 2000s onward, the number of CVD-related deaths increased steadily in the Midwest, North, and Northeast regions, whereas the Southeast and South regions maintained consistently high but relatively stable numbers of deaths (Figures 1B and 1C). The overall mortality rate also increased over the study period, rising from 7.3% in 1998 to 8.3% in 2023, with a steeper upward trend after 2012 and a peak of approximately 10.1% in 2021 (Figure 1D). Marked changes were also observed in the regional distribution of CVD mortality rates over time. In 2008, the Southeast region exhibited the highest mortality rate, whereas by 2022 the Midwest ranked first, with the North and Northeast reaching similarly elevated levels. By 2023, the Northeast had become the region with the highest CVD mortality rate, while the South consistently exhibited the lowest rates from 2008 onward (Figures 1E and 1F).

Based on this analysis, we focused on the major diseases directly affecting the cardiovascular system, selecting those responsible for the greatest mortality burden: chronic rheumatic heart diseases (I05–I09), hypertensive diseases (I10, I13–I15), hypertensive heart disease (I11), acute myocardial infarction (I21), chronic ischaemic heart disease (I25), cardiomyopathies (I42), and heart failure (I50) (Figures 2A and 2B). Cerebrovascular diseases (e.g., stroke) and other circulatory disorders were not included in the subsequent analyses, despite their clinical importance, because the present study aimed to characterise the epidemiology of disorders primarily involving the heart and cardiovascular system. Among the selected conditions, acute myocardial infarction (I21) remained the leading cause of death throughout the study period. Heart failure (I50) ranked second until 2016, after which it was overtaken by hypertensive diseases (I10, I13–I15), which remained the second leading cause of mortality through 2023 (Figure 2B). This shift occurred at different times across Brazilian regions, emerging earlier in the Northeast (around 2007) and later in the remaining regions, after 2014 (Figure 2C).

**Figure 2.**
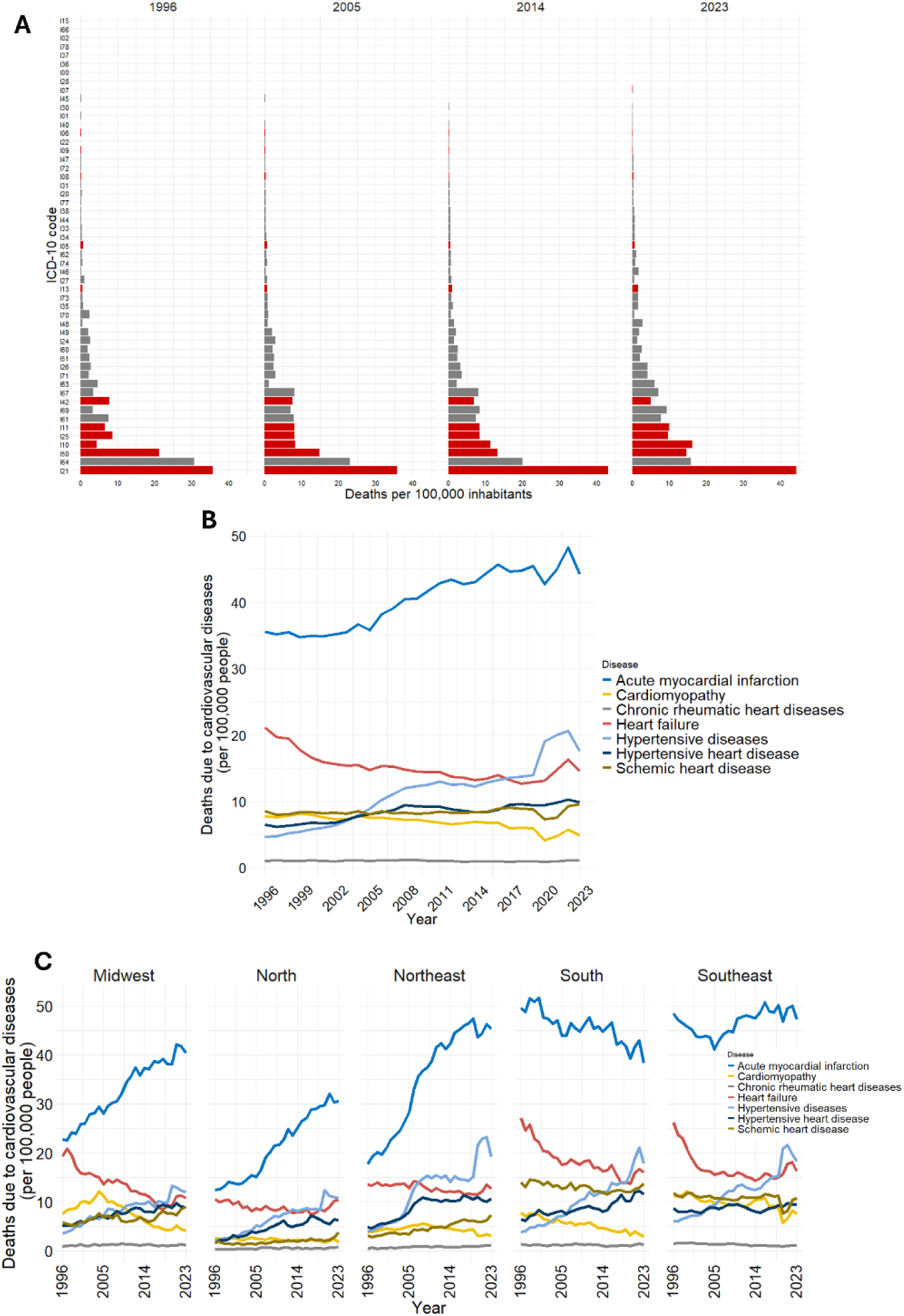
Temporal evolution of the leading causes of cardiovascular disease (CVD) mortality in Brazil (1996–2023). (**a**) Data was collected from the DATASUS platform and the classification was made according to the 10th revision of the International Classification of Diseases (ICD-10), Chapter IX Diseases of the circulatory system. Each code indicates a specific disease, as follows: I15 Secondary hypertension, I02 Rheumatic chorea, I66 Occlusion and stenosis of cerebral arteries not resulting in cerebral infarction, I78 Diseases of the capillaries, I37 Disorders of the pulmonary valve, I36 Non-rheumatic disorders of the tricuspid valve, I00 Rheumatic fever without mention of cardiac involvement, I28 Other diseases of the pulmonary vessels, I07 Rheumatic diseases of the tricuspid valve, I45 Other conduction disorders, I30 Acute pericarditis, I01 Rheumatic fever with cardiac involvement, I40 Acute myocarditis, I06 Rheumatic diseases of the aortic valve, I22 Recurrent myocardial infarction, I09 Other rheumatic diseases of the heart, I47 Paroxysmal tachycardia, I72 Other Aneurysms, I08 Multiple valve diseases, I31 Other pericardial diseases, I20 Angina pectoris, I77 Other diseases of arteries and arterioles, I38 Unspecified valve endocarditis, I44 Atrioventricular and left bundle branch block, I33 Acute and subacute endocarditis, I34 Non-rheumatic mitral valve disorders, I05 Rheumatic mitral valve diseases, I62 Other non-traumatic intracranial hemorrhages, I74 Arterial embolism and thrombosis, I46 Cardiac arrest, I27 Other forms of pulmonary heart disease, I13 Hypertensive heart and kidney disease, I73 Other peripheral vascular diseases, I35 Non-rheumatic aortic valve disorders, I70 Atherosclerosis, I48 Atrial flutter and fibrillation, I12 Hypertensive renal disease, I49 Other cardiac arrhythmias, I24 Other acute ischemic heart disease, I60 Subarachnoid hemorrhage, I51 Complications of heart disease and poorly defined heart conditions, I26 Pulmonary embolism, I71 Aortic aneurysm and dissection, I63 Cerebral infarction, I67 Other cerebrovascular diseases, I42 Cardiomyopathies, I69 Sequelae of cerebrovascular diseases, I61 Intracerebral hemorrhage, I11 Hypertensive heart disease, I25 Chronic ischemic heart disease, I10 Essential (primary) hypertension, I50 Heart failure, I64 Stroke, not specified as hemorrhagic or ischemic, and I21 Acute myocardial infarction. The graphs show the relationship between them every nine years. Only the diseases highlighted in red, corresponding to the most prevalent causes of CVD mortality, are further analysed and discussed in the manuscript. (**b**) Timeline of the seven most common causes of CVD death per 100,000 inhabitants, from 1996 to 2023. Acute myocardial infarction (I21) and hypertensive heart disease (I21) have become the most common causes of CVD death. Heart failure (I50) remains a concerning cause of death. (**c**) Number of deaths from the seven most common CVDs in Brazil per 100,000 inhabitants varies according to geographic region. All regions show an increase in mortality from acute myocardial infarction (I21), which is the most common cause of CVD death in all regions.

Regarding sex-specific mortality, acute myocardial infarction remained the leading cause of death in both men and women between 1996 and 2023, with mortality rates consistently higher in men (approximately 21.1–26.5 deaths per 100,000 inhabitants) than in women (14.6–17.6 deaths per 100,000 inhabitants). Heart failure and hypertensive diseases were the second and third leading causes of death, respectively. Heart failure mortality declined steadily in both sexes, decreasing from approximately 10 to 7.5 deaths per 100,000 inhabitants over the study period. In contrast, mortality from hypertensive diseases increased markedly, rising from about 3 to nearly 10 deaths per 100,000 inhabitants, with consistently higher rates among women than men, particularly in the later years of the series. Records with unavailable sex information remained consistently low throughout the study period (Figure 3A).

**Figure 3.**
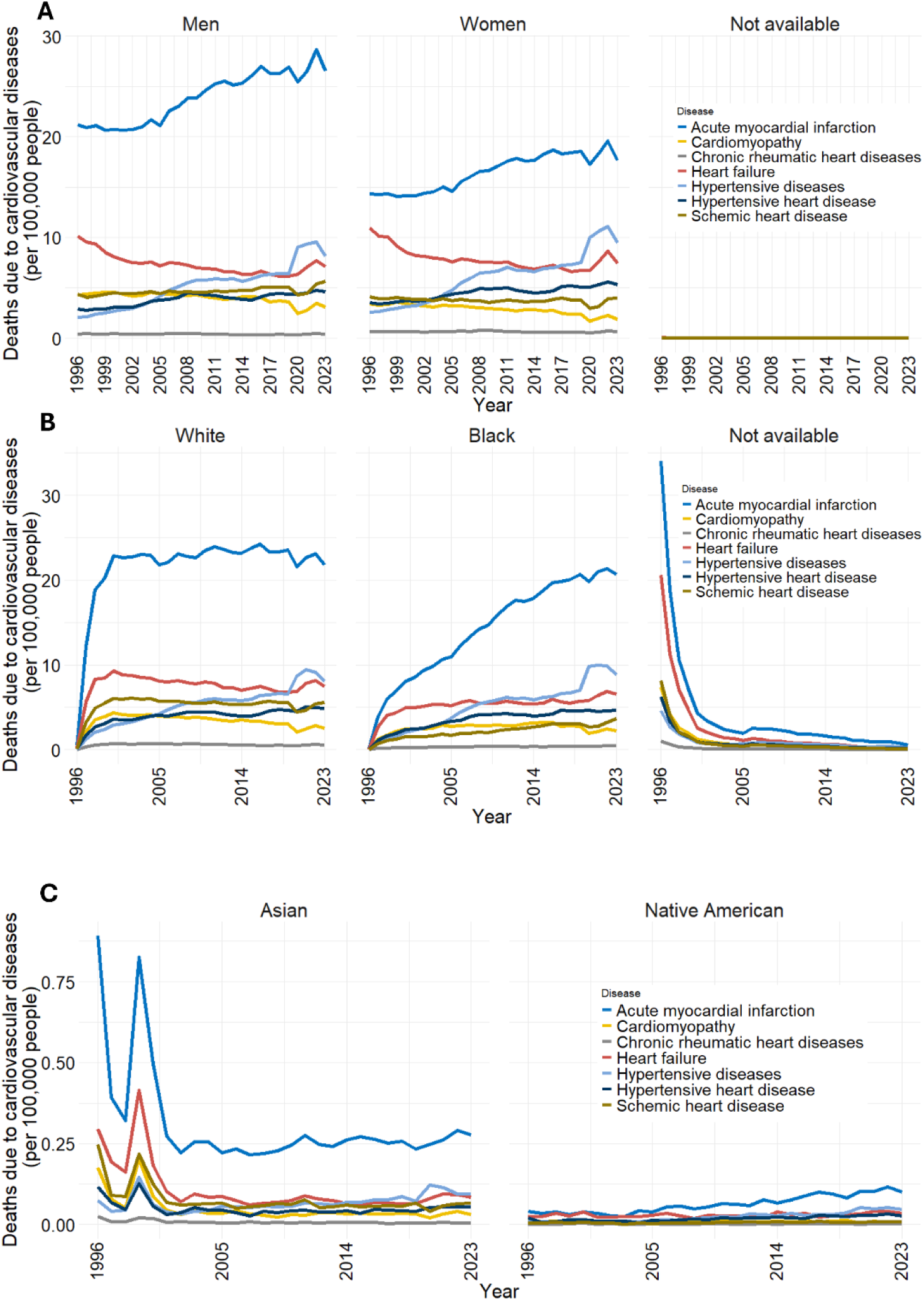
Profile of selected cardiovascular diseases (CVD) according to sex and ethnicity in Brazil. (**a**) Temporal trends in mortality from the seven selected CVDs according to sex. Acute myocardial infarction remained the leading cause of death in both sexes and consistently accounted for substantially more deaths in men than in women, whereas the remaining CVDs showed smaller sex-related differences. (**b**) Temporal trends in mortality according to self-reported ethnicity (White, Black, and Not available). Acute myocardial infarction was the leading cause of death in all groups, while hypertensive diseases contributed proportionally more to mortality among Black individuals. (**c**) Temporal trends in mortality among individuals of Asian descent and Indigenous populations. Mortality rates were considerably lower than those observed in the other ethnic groups, although acute myocardial infarction remained the predominant cause of death. Mortality rates are expressed as deaths per 100,000 inhabitants.

Regarding racial differences in CVD mortality, white and black populations exhibited distinct temporal patterns. Among white individuals, mortality from acute myocardial infarction increased sharply during the late 1990s and subsequently remained relatively stable at high levels, whereas among black individuals it showed a gradual and sustained increase throughout the study period. In both populations, mortality from hypertensive diseases progressively increased, whereas heart failure mortality declined only among white individuals. Consequently, hypertensive diseases overtook heart failure as the second leading cause of death earlier in the black population and later among whites. The proportion of records classified as “not available” decreased markedly over time, likely reflecting improvements in death certificate reporting (Figure 3B).

Among Asian populations, acute myocardial infarction also emerged as the leading cause of death, although mortality estimates were more variable during the early years of the study. The ranking of the remaining diseases was less consistent, with hypertensive diseases becoming the second leading cause toward the end of the study period (Figure 3C). Mortality data for Native American populations were highly variable throughout the series, likely owing to the small number of recorded deaths, although acute myocardial infarction became the predominant cause of death in the later years (Figure 3C).

We also investigated the impact of CVD across age groups. CVD-related mortality remained low among individuals younger than 50 years, increased progressively from 50–59 years onward, and reached its highest levels among those aged 80 years or older. Despite this age-dependent pattern, overall CVD mortality declined across all age groups between 1996 and 2023, with the largest reduction observed in individuals aged 80+ years (Figure 4A).

**Figure 4.**
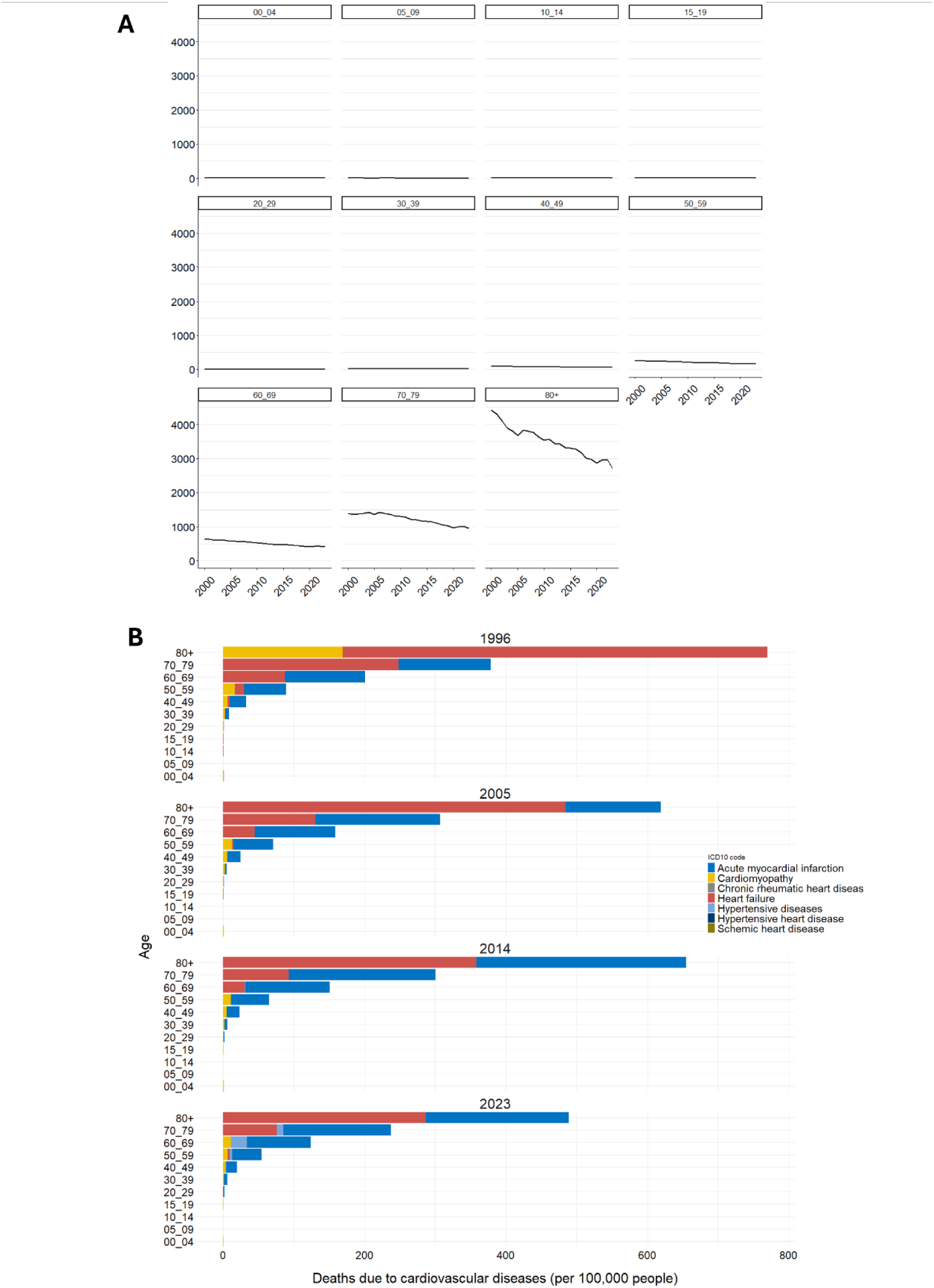
Age-specific patterns of cardiovascular disease (CVD) mortality in Brazil. (**a**) Temporal trends in age-specific CVD mortality rates from 1996 to 2023. Mortality increased markedly with advancing age, particularly after 50 years, while remaining relatively stable over time within each age group. The ≥80-year group exhibited the highest mortality rates throughout the study period, despite a gradual decline over time. Lines represent the mean mortality rate and shaded areas indicate the 95% confidence interval. (**b**) Distribution of the leading causes of CVD death according to age group in 1996, 2005, 2014, and 2023. Acute myocardial infarction was the leading cause of death from 30 years of age onward, whereas heart failure predominated among individuals aged ≥80 years throughout the analysed period.

When analysing causes of death across age groups in 1996, heart failure was the leading cause among individuals aged 70 years or older, whereas acute myocardial infarction was the leading cause among those aged 30–69 years. The second leading cause varied across age groups: cardiomyopathy ranked second among individuals aged 80 years or older and those younger than 60 years, whereas acute myocardial infarction ranked second among individuals aged 60–79 years (Figure 4B).

By 2005, acute myocardial infarction had become the leading cause of death among individuals aged 40–79 years and had increased substantially among those aged 80 years or older, becoming the second leading cause of death in this age group. By 2014, it accounted for nearly half of CVD-related deaths among individuals aged 80 years or older. In 2023, it remained the leading cause of death among individuals aged 30–79 years and the second leading cause among those aged 80 years or older. Heart failure remained the leading cause of death in the oldest age group throughout the study period (Figure 4B).

Importantly, from 1996 to 2023, both disease-specific and overall CVD mortality rates declined across age groups, particularly among older individuals. Among individuals aged 80+ years, mortality rates decreased from nearly 800 deaths per 100,000 inhabitants in 1996 to less than 500 per 100,000 in 2023. Similarly, among individuals aged 70–79 years, mortality rates declined from nearly 400 deaths per 100,000 inhabitants in 1996 to less than 250 per 100,000 in 2023 (Figure 4B).

### The economic burden of cardiovascular diseases in Brazil

The overall expenditure associated with CVD-related hospitalizations increased in Brazil between 1998 and 2023 (Figure 5A), although the magnitude of this increase varied considerably across its regions (Figure 5B). The Southeast region consistently accounted for the highest hospitalization costs, increasing from approximately BRL 1.45 billion (≈ USD 290 million) in 1998 to more than BRL 2,10 billion (≈ USD 420 million) in 2023. Similarly, expenditures in the South and Northeast regions increased from BRL 640.9 million (≈ USD 128 million) to BRL 1.09 billion (≈ USD 220 million), and from BRL 487.8 million (≈ USD 98 million) to BRL 900.9 million (≈ USD 180 million), respectively. Despite presenting the lowest absolute cost compared with the other regions, the North region showed the highest increase, with expenditures rising approximately 3-fold between 1998 and 2023 (Figure 5B).

**Figure 5.**
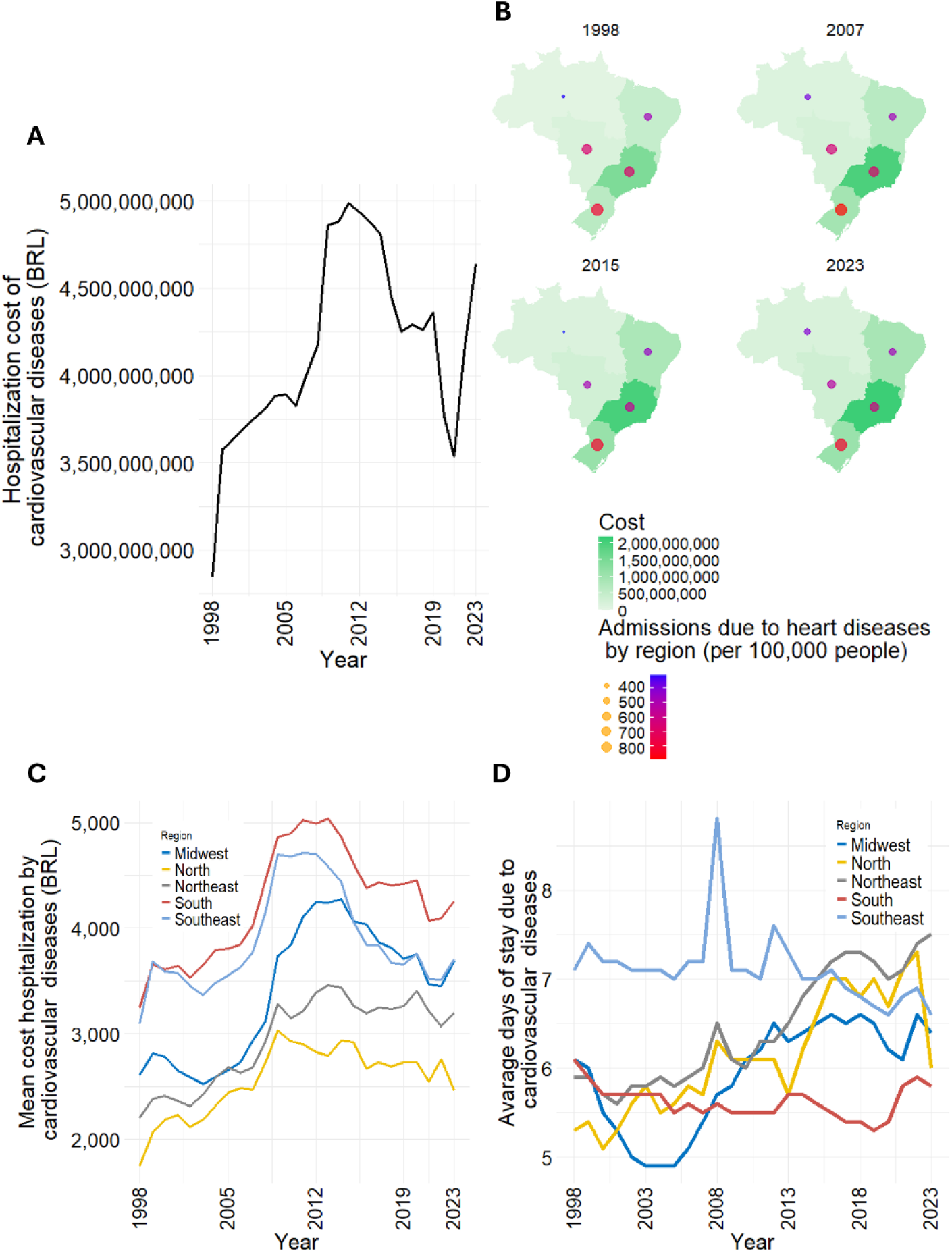
Hospitalizations and healthcare costs associated with cardiovascular diseases (CVD) in Brazil. (**a**) Total annual expenditure on CVD-related hospitalizations from 1998 to 2023. Values were adjusted for inflation using the Extended National Consumer Price Index (Índice Nacional de Preços ao Consumidor Amplo, IPCA) calculated by the Brazilian Institute of Geography and Statistics (IBGE). (**b**) Regional trends in total CVD-related hospitalization expenditure and hospital admissions per 100,000 inhabitants from 1998 to 2023. (**c**) Mean cost per CVD-related hospitalization according to Brazil’s geographic regions between 1998 and 2023. (**d**) Average length of CVD-related hospital stay according to Brazil’s geographic regions between 1998 and 2023.

CVD-related hospital admissions per 100,000 inhabitants displayed distinct regional patterns. The Midwest, Northeast and Southeast regions showed an initial reduction up to 2015, followed by a modest increase in 2023. The South region had an early increase up to 2007, followed by a slight decline, reaching rates similar to 1998. The North region showed a continuous increase throughout the period, rising from 322.3 to 417.7 admissions per 100,000 inhabitants from 1998 to 2023 (Figure 5B). Detailed CVD-related hospitalization costs and admission rates for each Brazilian region are presented in Table S5 (supplemental material).

All five regions showed a similar trend in the mean cost of hospitalization, with costs increasing until the early 2010s, when they reached their peak values, followed by a gradual decline through 2023. The South and Southeast regions consistently exhibited the highest mean hospitalization costs throughout the study period, whereas the North region showed the lowest values. The Midwest experienced a marked increase between 2005 and 2013, reaching levels comparable to those observed in the Southeast (Figure 5C).

The average length of CVD-related hospital stay remained relatively stable across most Brazilian regions throughout the analysed period (Figure 5D). The North and Northeast regions were exceptions, showing a gradual increase over time. In the North, the average stay rose from approximately 5.4 days in 1998 to 6.0 days in 2023, peaking at 7.3 days in 2022, whereas in the Northeast it increased steadily from 5.8 to 7.5 days over the same period. The South region remained comparatively stable, fluctuating between 5.3 and 6.0 days and exhibiting the shortest hospital stays by 2023. The Midwest showed an initial decline followed by a gradual increase after 2010, whereas the Southeast consistently presented the longest hospital stays until around 2016, when it was surpassed by the Northeast, which thereafter exhibited the longest average length of stay among all Brazilian regions.

### Heart transplant panorama in Brazil

We analysed the impact of heart transplantation on Brazil’s public health system (SUS), as it represents a last-line therapy for patients with severe, treatment-refractory heart disease (Figure 6). It is important to note that the unusually low number of heart transplants recorded in 2007 should be interpreted with caution, as this period coincided with the implementation of a new DATASUS procedure coding system and the unification of hospital and outpatient information databases, which likely affected data reporting and comparability during the transition. Since then, the number of heart transplants progressively increased, reaching a peak of 0.162 transplants per 100,000 inhabitants in 2019 and remaining at similarly high levels in 2023 (0.136 per 100,000 inhabitants) (Figure 6A). The number of transplantation-related intercurrences also increased over time, rising from fewer than 0.05 cases per 100,000 inhabitants in 2012 to approximately 0.4 in 2023, when it reached its highest level (Figure 6B).

**Figure 6.**
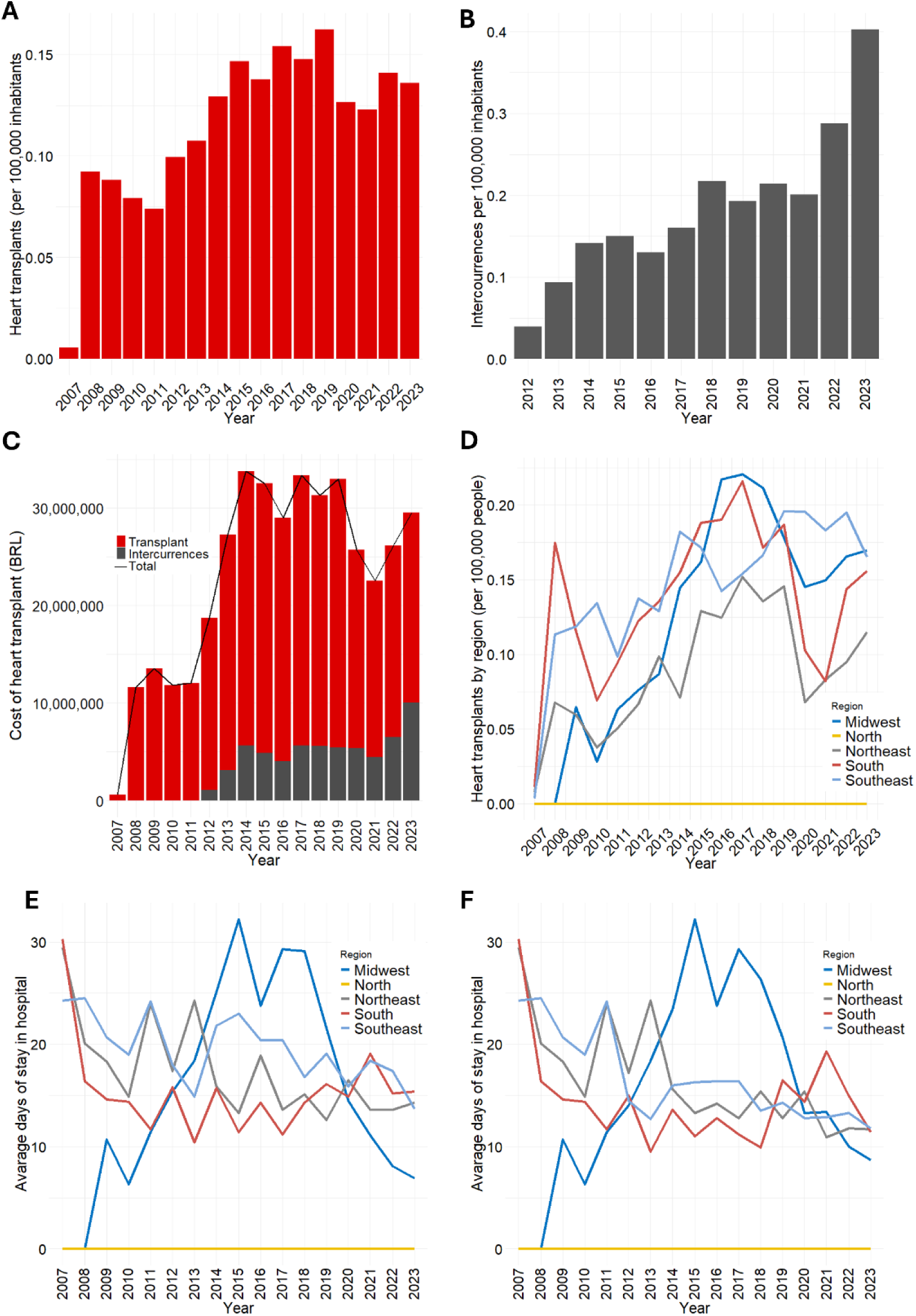
Heart transplantation in Brazil from 2007 to 2023. (**a**) Annual number of heart transplants per 100,000 inhabitants in Brazil. (**b**) Annual number of heart transplantation-related intercurrences per 100,000 inhabitants from 2012 to 2023. (**c**) Total annual expenditure on heart transplantation in Brazil (BRL). Values were adjusted for inflation using the Extended National Consumer Price Index (“Índice Nacional de Preços ao Consumidor Amplo”, IPCA) calculated by the Brazilian Institute of Geography and Statistics (IBGE). (**d**) Heart transplantation rate (per 100,000 inhabitants) according to Brazil’s geographic regions from 2007 to 2023. (**e**) Average length of hospital stay following heart transplantation according to geographic region. (**f**) Average length of hospital stay associated with heart transplantation-related intercurrences according to geographic region.

The total expenditure on heart transplantation peaked in 2014, reaching BRL 33,807,575.20 (≈ USD 6.7 million). Costs remained relatively high until 2020, when a temporary decline was observed, before increasing again through 2023 (Figure 6C).

Between 2007 and 2023, heart transplantation rates generally increased across the Brazilian regions, although substantial year-to-year fluctuations were observed. The Midwest, South, and Northeast regions reached their highest rates between 2016 and 2018, followed by a decline during 2020–2021. In contrast, the Southeast peaked around 2018–2019 and maintained relatively high transplantation rates thereafter. Notably, the North region recorded no heart transplants throughout the study period (Figure 6D).

The average length of hospitalization for heart transplantation (Figure 6E) and transplantation-related intercurrences (Figure 6F) showed very similar regional patterns throughout the analysed period. The South, Southeast, and Northeast regions exhibited an overall decline in hospitalization duration, although with substantial year-to-year fluctuations. In the South and Northeast, the average length of stay decreased from approximately 30 days in 2007 to around 15 days by 2023, whereas the Southeast declined from approximately 25 to 15 days over the same period. In contrast, the Midwest displayed a distinct pattern, with hospitalization duration increasing sharply between 2010 and 2015, peaking at more than 30 days, before declining to fewer than 10 days by 2023. As expected, the North region consistently recorded zero hospitalization days due to the absence of heart transplant procedures during the study period (Figures 6E and 6F).

## Discussion

The epidemiological profile of Brazil has changed dramatically over the past three decades. In the early 1990s, communicable diseases (i.e., diarrhoeal diseases) were the leading causes of mortality ^12^, largely reflecting inadequate sanitation and the early stages of implementation of the Unified Health System (SUS) ^13,14^. Since then, Brazil has undergone a marked epidemiological transition, with CVD progressively becoming the leading cause of death ^12^. In this context, our study provides the most comprehensive longitudinal assessment of the CVD burden in Brazil encompassing nearly three decades (1996–2023). By integrating mortality, hospitalizations, regional disparities, ethnicity, age, sex, healthcare costs, and heart transplantation data, this study substantially expands a previous DATASUS-based report focused primarily on CVD-related emergency hospitalizations between 2013 and 2023 ^15^, providing a more comprehensive perspective on the epidemiological and economic burden of CVD in Brazil and its implications for public health planning and resource allocation. This long-term analysis also allowed the identification of important temporal disruptions in CVD indicators, particularly during the COVID-19 pandemic. A marked reduction in several indicators was observed in 2020, likely reflecting disruptions in health information systems and changes in healthcare-seeking behaviour during the COVID-19 pandemic. The subsequent rebound may also reflect the indirect cardiovascular consequences of delayed diagnosis and treatment during this period ^16–18^. Although CVD-related healthcare expenditures increased again after 2020, they did not return to the peak levels observed before the pandemic, suggesting a persistent impact on healthcare utilisation and resource allocation.

Between 1996 and 2023, CVD deaths in Brazil (Human Development Index [HDI] = 0.805) increased, reaching 383,474 deaths in 2023, approximately 180.0 per 100,000 inhabitants. By comparison, the United States (HDI = 0.938) reported 129.5 deaths per 100,000 inhabitants in 2022, France (HDI = 0.920) 62.6, whereas Uzbekistan (HDI = 0.740) recorded the highest rate worldwide in 2023, with 594.1 deaths per 100,000 inhabitants. Although multiple factors contribute to these differences, these comparisons are consistent with an inverse association between socioeconomic development and CVD mortality, which has been reported globally ^19,20^ and within Brazilian cities ^21^.

Regional disparities were among the most important findings of this study. The Southeast and South regions consistently recorded the highest numbers of CVD-related deaths, probably reflecting their larger and older populations ^22^. However, when mortality rates were considered, the Northeast region exhibited the highest CVD mortality rate in 2023. Nonetheless, lower mortality rates observed in other regions should be interpreted cautiously, as they may partly reflect underdiagnosis and underreporting. Evidence from municipalities in the Amazon region has demonstrated substantial discrepancies between local notification systems and the national DATASUS database, exposing important limitations in disease registration and surveillance ^23^. This issue is particularly relevant for the North region, which simultaneously showed the largest increase in CVD-related hospitalizations yet recorded no heart transplants throughout the study period. Together with its lower mortality rates, these findings highlight persistent inequalities in access to highly specialised cardiovascular care and suggest that both healthcare availability and information system limitations require greater attention in this region.

The Brazilian Institute of Geography and Statistics (IBGE) classifies ethnicity into five groups (Black, Brown, Asian, Indigenous, and White) based on self-declared skin colour. In epidemiological studies, Black and Brown populations are commonly combined to represent the Black population, an approach adopted here. Within this framework, hypertensive disease mortality increased disproportionately among Black individuals. Although genetic susceptibility has been proposed, biological factors alone cannot explain these disparities. Structural racism, unequal access to healthcare, delayed diagnosis, differences in treatment, and socioeconomic inequalities likely contribute substantially to these outcomes ^24–27^.

Similarly, the markedly lower number of CVD-related cases observed among Indigenous populations is likely influenced by underreporting, a limitation previously described in other epidemiological studies in Brazil ^28,29^. Geographic isolation, barriers to healthcare access, and limited health infrastructure may contribute to incomplete case registration, suggesting that the true burden of CVD in these communities is substantially underestimated.

Sex and age emerged as major determinants of CVD mortality in the present study, corroborating previous evidence that both are major determinants of cardiovascular risk and mortality ^30,31^. Acute myocardial infarction consistently accounted for substantially more deaths in men than in women, whereas sex differences for other CVDs were less pronounced. This male predominance may reflect both greater exposure to behavioural risk factors and an intrinsically faster biological ageing process, which has recently been shown to explain a substantial proportion of the excess cardiovascular risk observed in men, independent of traditional risk factors and sex hormones ^32^. Age had an even greater influence on CVD mortality, with death rates increasing exponentially after 50 years of age. Heart failure predominated among the oldest individuals, whereas acute myocardial infarction became the leading cause of death from 30 years of age onward, highlighting distinct age-related patterns in the burden of CVD.

## Conclusion

CVD remains a leading cause of death in Brazil and worldwide, imposing heavy epidemiological and economic burdens. Mortality rates stay high across Brazil, with disparities by time, region, ethnicity, sex, and age. Acute myocardial infarction dominates, while hypertension and heart failure also contribute significantly. The study highlights rising hospitalization and transplant costs, regional inequalities in access and reporting, and the urgent need for stronger surveillance, equitable care, and preventive strategies targeting vulnerable groups and modifiable risks.

## Data Availability

All data produced in the present study are available upon reasonable request to the authors

## Contributors

AGO and TVB conceptualized the idea of the analysis and research, with input from all authors. AGO and TVB were responsible for funding acquisition, determining the methodology for this study and project administration. MCBS and JSFG performed data curation and investigation of all the data in this study, with input from ALMO, AGO and TVB. MCBS, JSFG, TVB, and AGO were responsible for the formal analysis with contributions from all authors. ALMO conducted data curation for correction of costs based on the national extended consumer price index throughout the years. MCBS, JSFG and TVB have directly accessed and validated the data. MCBS, JSFG were responsible for writing the original draft, with significant contributions from all authors. MCBS, JSFG TVB, and AGO were responsible for writing-reviewing and editing the final version of this draft, with significant contributions from all authors.

## Data sharing statement

The data used for this study were retrieved from public online repositories maintained by the Brazilian Government. The system allows anyone who wishes to access the data (https://datasus.saude.gov.br/informacoes-de-saude-tabnet/). Data can be retrieved from different sets as follows: Morbidity (https://datasus.saude.gov.br/acesso-a-informacao/morbidade-hospitalar-do-sus-sih-sus/), Mortality (http://tabnet.datasus.gov.br/cgi/deftohtm.exe?sim/cnv/obt10uf.def), and Transplants (http://tabnet.datasus.gov.br/cgi/deftohtm.exe?sih/cnv/qruf.def).

## Acknowledgments and Disclaimer

This work was supported by CNPq (Scholarship to JSFG and TVB’s grants: 404407/2025-0, 444243/2024-0, 403725/2024-0, 406936/2023-4, and 309965/2022-5), DAAD/CAPES (TVB: 88881.700905/2022-01), CAPES - Finance Code 001 (Scholarship to MCBS and TVB’s grant: 88887.916694/2023-00), FAPEMIG (TVB: BPD-00133-22), and INCT-Hepatologia 360 (AGO: INCT 407909/2024-9).

The views expressed in this document are those of the authors and do not necessarily reflect the views of the United Nations or the countries it represents.

---

**Supplementary Table S1:** Heart diseases defined by the International Classification of Diseases (ICD10)

| Code | ICD-10 Category |
| --- | --- |
| I00 | Rheumatic fever without mention of heart involvement |
| I01 | Rheumatic fever with heart involvement |
| I02 | Rheumatic chorea |
| I05 | Rheumatic mitral valve diseases |
| I06 | Rheumatic aortic valve diseases |
| I07 | Rheumatic tricuspid valve diseases |
| I08 | Multiple valve diseases |
| I09 | Other rheumatic heart diseases |
| I10 | Essential (primary) hypertension |
| I11 | Hypertensive heart disease |
| I13 | Hypertensive heart and renal disease |
| I15 | Secondary hypertension |
| I20 | Angina pectoris |
| I21 | Acute myocardial infarction |
| I22 | Subsequent myocardial infarction |
| I23 | Certain current complications following acute myocardial infarction |
| I24 | Certain current complications following acute myocardial infarction |
| I25 | Chronic ischaemic heart disease |
| I26 | Pulmonary embolism |
| I27 | Other pulmonary heart diseases |
| I28 | Other diseases of pulmonary vessels |
| I30 | Acute pericarditis |
| I31 | Other diseases of pericardium |
| I33 | Acute and subacute endocarditis |
| I34 | Nonrheumatic mitral valve disorders |
| I35 | Nonrheumatic aortic valve disorders |
| I36 | Nonrheumatic tricuspid valve disorders |
| I37 | Pulmonary valve disorders |
| I38 | Endocarditis, valve unspecified |
| I40 | Acute myocarditis |
| I42 | Cardiomyopathy |
| I44 | Atrioventricular and left bundle-branch block |
| I45 | Other conduction disorders |
| I46 | Cardiac arrest |
| I47 | Paroxysmal tachycardia |
| I48 | Atrial fibrillation and flutter |
| I49 | Other cardiac arrhythmias |
| I50 | Heart failure |
| I51 | Complications and ill-defined descriptions of heart disease |
| I60 | Subarachnoid haemorrhage |
| I61 | Intracerebral haemorrhage |
| I62 | Other nontraumatic intracranial haemorrhage |
| I63 | Cerebral infarction |
| I64 | Stroke, not specified as haemorrhage or infarction |
| I66 | Occlusion and stenosis of cerebral arteries, not resulting in cerebral infarction |
| I67 | Other cerebrovascular diseases |
| I69 | Sequelae of cerebrovascular disease |
| I70 | Atherosclerosis |
| I71 | Aortic aneurysm and dissection |
| I72 | Other aneurysm and dissection |
| I73 | Other peripheral vascular diseases |
| I74 | Arterial embolism and thrombosis |
| I77 | Other disorders of arteries and arterioles |
| I78 | Diseases of capillaries |

**Supplementary Table S2:** Description of variables analysed.

| Variable | Description |
| --- | --- |
| Sex | Sex Sex of the patient (male, female) |
| Race/ethnicity | Race/ethnicity of the patient (White, Black, Brown, Asian, Indigenous, or unspecified) |
| ICD10 chapter | Cause of hospitalization, according to the International Classification of Diseases (ICD),retrieved by chapters. |
| ICD10 list | Cause of hospitalization, according to the International Classification of Diseases (ICD),retrieved by list. |
| ICD10 Category | Cause of hospitalization, according to the International Classification of Diseases (ICD),retrieved by category . |
| Age group | Under 1 year, 1–4 years, 5–9 years, 10–14 years, 15–19 years, 20–29 years, 30–39 years, 40–49 years, 50–59 years, 60–69 years, 70–79 years, 80 years and older. |

**Supplementary Table S3:** Description of parameters analysed.

| Parameters | Description |
| --- | --- |
| Hospitalization admission | Number of HAA (Hospital Admission Authorization) approved during the period, excluding extensions (long-stay cases). This is an approximate value of hospitalizations, as transfers and readmissions are included in this count |
| Total hospitalization cost | The value referring to the approved HAA (Hospital Admission Authorization) during the period. This value may not necessarily correspond to the amount transferred to the healthcare facility, as it depends on the situation of the units. They may receive budgetary resources or there may be retentions and payments of incentives, which are not presented here. Therefore, this value should be considered as the approved production value |
| Mean cost of hospitalization admission | Total hospitalization cost divided by the number of hospitalizations |
| Average days of stay in hospital | Average length of stay for hospitalizations related to approved HAA (Hospital Admission Authorization), recorded as admissions, during the period. |
| Deaths | Number of hospitalizations that resulted in discharge due to death among the approved HAA (Hospital Admission Authorization) during the period. |
| Mortality rate | The ratio between the number of deaths and the number of approved HAA (Hospital Admission Authorization) recorded as hospitalizations during the period, multiplied by 100. |

**Supplementary Table S4:** Hospital Morbidity in the Unified Health System of Brazil.

| Chapter | Code | Description | ICD10 |
| --- | --- | --- | --- |
| IX | 066-072 | Diseases of the circulatory system | I00-I99 |

**Supplementary Table S5:** Relative admissions and hospitalisation cost by regions.

| Year | Region | Admissions (per 100,000) | Cost (in BRL) |
| --- | --- | --- | --- |
| 1998 | Midwest | 672.7167 | 192,959,555 |
| 1998 | North | 322.2924 | 66,846,381 |
| 1998 | Northeast | 482.8149 | 487,769,710 |
| 1998 | South | 817.8246 | 640,926,201 |
| 1998 | Southeast | 681.7196 | 1,453,770,982 |
| 2007 | Midwest | 645.6512 | 250,704,331 |
| 2007 | North | 395.4090 | 143,000,080 |
| 2007 | Northeast | 483.5087 | 669,003,818 |
| 2007 | South | 893.0429 | 960,454,497 |
| 2007 | Southeast | 675.8243 | 1,986,099,584 |
| 2015 | Midwest | 493.9312 | 309,871,057 |
| 2015 | North | 316.3253 | 161,921,327 |
| 2015 | Northeast | 456.1804 | 843,431,002 |
| 2015 | South | 827.7951 | 1,117,167,726 |
| 2015 | Southeast | 579.4985 | 2,018,729,278 |
| 2023 | Midwest | 541.0276 | 341,468,367 |
| 2023 | North | 417.6548 | 199,353,135 |
| 2023 | Northeast | 483.4854 | 900,931,365 |
| 2023 | South | 829.4803 | 1,086,713,935 |
| 2023 | Southeast | 627.2055 | 2,108,607,004 |

